# Trauma in healthcare staff: A multiple methods study using quantitative and qualitative lived experience of participants in a randomised controlled trial of a brief digital imagery-competing task intervention for intrusive memories

**DOI:** 10.64898/2026.03.02.26347416

**Authors:** Priya Patel, Susan Brown, Alfred Markham, Amy Beckenstrom, Maja Friedemann, Jonathan Kingslake, Julie Highfield, Charlotte Summers, Emily A Holmes, Richard Morriss

## Abstract

**Objective:** This mixed-methods study investigated the lived-experience perspectives of receiving a novel, brief digital mental health intervention after psychological trauma. The online gamified imagery-competing task intervention (ICTI) involves one researcher-guided session followed by self-use. Tested in two randomised controlled trials (GAINS-01; GAINS-02), ICTI led to fewer intrusive memories at week-4, with the reduction sustained over 24 weeks, alongside reductions in post-traumatic stress. Here, we contrasted user experiences of ICTI with an Active Control (AC; music-listening task), and explored longer-term impact in qualitative interviews to contextualise GAINS-02 findings.

**Methods and Analysis:** The GAINS-02 trial randomised healthcare staff experiencing intrusive memories after work-related trauma to ICTI (N=40), AC (N=39), or treatment-as-usual (TAU; N=20). Expectancy was assessed before the researcher-guided session (Day 0), acceptability at week-4, and usage tracked for 24-weeks. Semi-structured interviews (N=27) were conducted in ICTI and AC arms only (15 at week-4; 12 during 12-24-weeks). Interviews were analysed using reflexive thematic analysis.

**Results:** Prior to use, many trial participants did not think the intervention would work, favouring AC over ICTI. However, after completing the tasks, participants found ICTI more acceptable and relevant to intrusive memories than AC. After the one guided session, median ICTI usage the next four weeks was 4.00 times with little additional use (once more) over the next 20 weeks because of lack of need.

Potential implementation facilitators included ease of use, and advantages over existing interventions due to not needing to talk about the trauma, brevity, and lesser resource commitment. Perceived barriers included a lack of staff and manager education about the nature and consequences of intrusive memories, with a need for workplace buy-in and demonstration of organisational benefits.

**Conclusion:** Healthcare staff experiencing workplace-related trauma found ICTI to be acceptable and effective for reducing intrusive memories with low effort and emotional burden, even among participants who initially expressed scepticism. Participants highlighted implementation considerations including offering ICTI both within and outside the workplace, and providing a self-guided version of ICTI with optional support. Future work should assess cost-effectiveness, impacts on presenteeism and retention, and real-world implementation including the feasibility and effectiveness of a self-guided ICTI.

**Summary Box:** *What is already known on this topic:* In a previous randomised controlled trial (GAINS-01) with Intensive Care Unit (ICU) staff exposed to work-related trauma, a brief online gamified imagery-competing task intervention (ICTI) reduced intrusive memories compared to usual care at four-weeks.

*What this study adds:* The GAINS-02 randomised controlled trial replicated GAINS-01 and extended results by comparing ICTI to an active control (AC; music listening) task, enrolling hospital staff from outside ICU, and a follow-up period of 24-weeks. Qualitative interviews found that, despite initial scepticism from healthcare staff prior to using the intervention, ICTI was more acceptable than an AC due to specific effects on swiftly reducing intrusive memories and requiring minimal support or usage after an initial researcher-guided session. After one guided session, ICTI was used 4 more times in the first four weeks, with little additional usage (once) thereafter because of lack of need (i.e., no longer experiencing intrusive memories).

*How this study might affect research, practice or policy:* ICTI is an efficacious scalable intervention to relieve staff of intrusive memories with effects sustained for at least 6-months. It was found to be more acceptable to participants than alternatives, requiring less time commitment than standard psychological treatments.

## 1. Introduction

Following the experience of psychological trauma, people may experience involuntary, sensory ’intrusive memories’, often as vivid visual imagery[1] triggered by reminders of the event. While these intrusions commonly subside over time, they can persist and form a core symptom of post-traumatic stress disorder (PTSD).[2]

Intrusive memories and other PTSD symptoms are a global issue among healthcare professionals exposed to work-related trauma,[3] including during the COVID-19 pandemic.[4] PTSD symptoms have been reported at elevated levels in NHS staff, particularly in intensive care settings, and are associated with adverse occupational outcomes such as sickness absence and intentions to leave.[5] Many healthcare professionals report limited access to adequate support.[6] NHS workforce plans[7] highlight the importance of effective health and wellbeing provision to support retention.

The imagery competing task intervention (ICTI) tested in the GAINS-01 and GAINS-02 randomised controlled trials (RCTs), is a brief online intervention during which participants briefly, intentionally bring to mind an intrusive memory image and then play Tetris® for 20-minutes while focusing on mental rotation.[8] ICTI was co-produced with healthcare staff with lived experienced of trauma[9] and optimised during development.[10]

In GAINS-01, [11, 12] intensive care staff were randomly assigned to ICTI or treatment as usual (TAU). Those receiving ICTI reported significantly fewer intrusive memories at week-4. A mixed-methods usability and acceptability study alongside GAINS-01[13] found that, despite initial scepticism, participants later rated ICTI as helpful, easy to use, and easier to fit into daily life than alternatives such as talking therapies. Participants valued that ICTI targets a specific symptom without requiring a formal diagnosis, and some highlighted the benefits of not needing to talk in detail about the initiating trauma. However, perceived barriers included stigma around mental ill-health, privacy concerns about the digital format, and practical constraints such as time. Findings highlighted the need to examine longer-term use and implementation.[13] An interview study with healthcare workers in Sweden[14] showed some convergent findings[13] but also raised questions about whether ICTI can be used without facilitator support, with implications for scalability.

GAINS-02[15] evaluated the optimised digital ICTI in a broader sample of NHS healthcare staff and with minimal facilitator input (a single online researcher-guided session and optional self-use thereafter, with optional researcher support for the first four-weeks). GAINS-02 was doubly controlled, comparing ICTI with an active control (AC; a 20-minute Mozart music-listening task delivered via the same platform and similar researcher contact time) and with treatment-as-usual (TAU), to assess whether effects could reflect non-specific or expectancy influences.[16, 17] Efficacy results are reported separately.[15]

Alongside GAINS-02 RCT, the current multiple-methods study aimed to: (1) compare treatment credibility and expectations of ICTI and AC before use; (2) assess feasibility and acceptability of ICTI at four-weeks; (3) investigate ICTI and AC usage across 24-weeks; (4) contrast experiences of ICTI and AC, and identify perceived benefits and challenges over the short and longer term, as examined via interviews (4-weeks, 12–24-weeks); (5) consider barriers and facilitators to scale-up and adoption of ICTI within routine NHS services; and (6) triangulate quantitative and qualitative findings to inform further development as ICTI moves towards implementation.

## 2. Materials and Methods

### 2.1 Patient and Public Involvement Statement

GAINS-01 and GAINS-02 trials[11, 13, 15] achieved meaningful involvement through a collaborative partnership with the Intensive Care Society (ICS) and tailored the study for ICU staff. From conception, the study team included members with lived experience of work-related trauma. ICS representatives contributed to refining the intervention and study materials, advised on recruitment strategies, and facilitated engagement via professional networks and social media within and beyond ICS. Ongoing weekly meetings with ICS representatives enabled continuous feedback, refinement, and shared decision making, and ICS partners were involved in dissemination of findings (including co-author JH).[8] This collaboration enabled the research to remain relevant, feasible, and grounded in the everyday work context. GAINS-02 further refined the intervention by incorporating feedback from GAINS-01 interview and stakeholder workshop participants, which included ICU and broader NHS staff.

### 2.2 Design

This multiple-methods study was conducted alongside the three-arm, parallel-group GAINS-02 RCT.[15] Participants were randomly allocated to either intervention (ICTI), active control (AC) or treatment-as-usual (TAU) (Supplementary Figure S1). ICTI consisted of access to the brief imagery-competing task intervention for 24-weeks. AC consisted of access to a brief music-listening task for 24-weeks. TAU consisted of usual care that participants would otherwise receive.

Data collected included: (i) Credibility and Expectancy Questionnaire (CEQ; Table 1; [18]), c (ii) task-usage data (iii) Acceptability Feedback Questionnaire (AFQ); Table 2); and (iv) qualitative interview data with a sub-sample at four-weeks and during 12-24-weeks (ICTI and AC; Supplementary Figure S1).

**Table 1.**
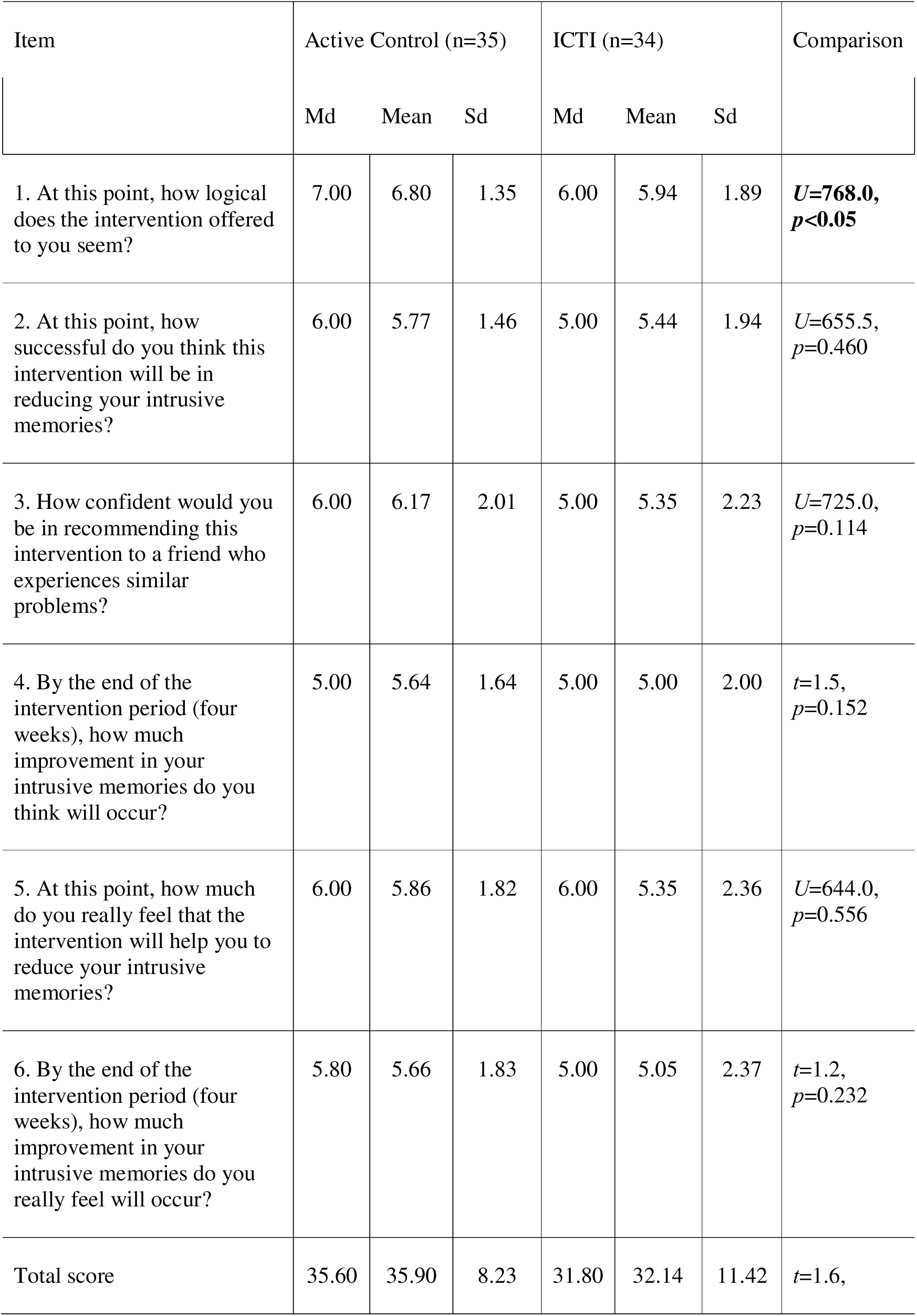
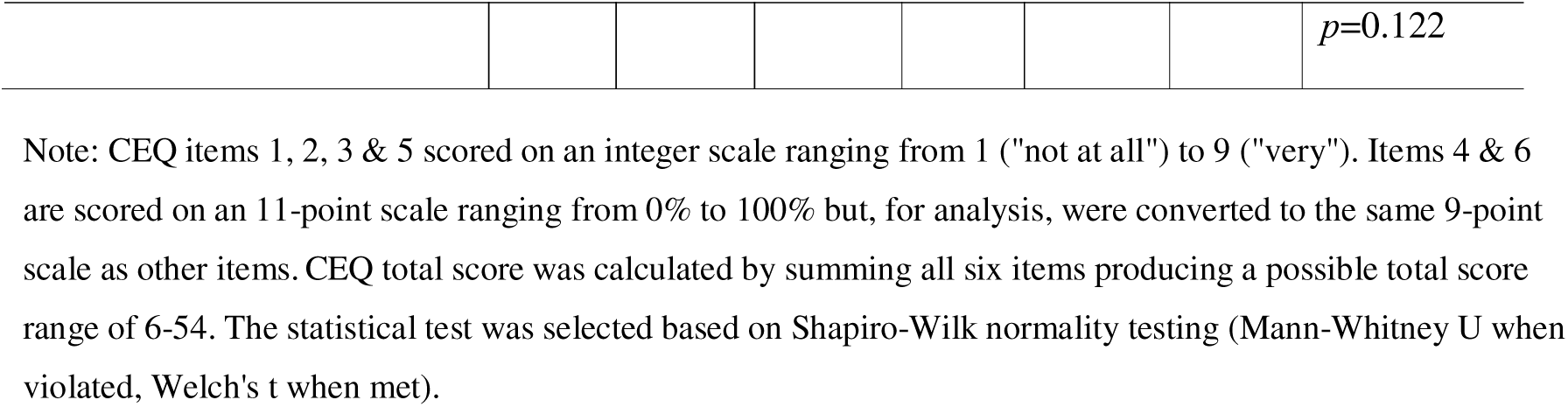
Credibility Expectancy Questionnaire (CEQ) per item at baseline (post-randomisation and before intervention use).

| Item | Active Control (n=35) |  |  | ICTI (n=34) |  |  | Comparison |
| --- | --- | --- | --- | --- | --- | --- | --- |
|  | Md | Mean | Sd | Md | Mean | Sd |  |
| 1. At this point, how logical does the intervention offered to you seem? | 7.00 | 6.80 | 1.35 | 6.00 | 5.94 | 1.89 | ***U=768.0, p<0.05*** |
| 2. At this point, how successful do you think this intervention will be in reducing your intrusive memories? | 6.00 | 5.77 | 1.46 | 5.00 | 5.44 | 1.94 | *U=655.5, p=0.460* |
| 3. How confident would you be in recommending this intervention to a friend who experiences similar problems? | 6.00 | 6.17 | 2.01 | 5.00 | 5.35 | 2.23 | *U=725.0, p=0.114* |
| 4. By the end of the intervention period (four weeks), how much improvement in your intrusive memories do you think will occur? | 5.00 | 5.64 | 1.64 | 5.00 | 5.00 | 2.00 | *t=1.5, p=0.152* |
| 5. At this point, how much do you really feel that the intervention will help you to reduce your intrusive memories? | 6.00 | 5.86 | 1.82 | 6.00 | 5.35 | 2.36 | *U=644.0, p=0.556* |
| 6. By the end of the intervention period (four weeks), how much improvement in your intrusive memories do you really feel will occur? | 5.80 | 5.66 | 1.83 | 5.00 | 5.05 | 2.37 | *t=1.2, p=0.232* |
| Total score | 35.60 | 35.90 | 8.23 | 31.80 | 32.14 | 11.42 | *t=1.6,* |
| | | | | | | | $p=0.122$ |

Note: CEQ items 1, 2, 3 & 5 scored on an integer scale ranging from 1 ("not at all") to 9 ("very"). Items 4 & 6 are scored on an 11-point scale ranging from 0% to 100% but, for analysis, were converted to the same 9-point scale as other items. CEQ total score was calculated by summing all six items producing a possible total score range of 6-54. The statistical test was selected based on Shapiro-Wilk normality testing (Mann-Whitney U when violated, Welch's t when met).

**Table 2.** Acceptability Feedback Questionnaire (AFQ) scores per item at 4-weeks.

| Item | Active Control (n=22) |  |  | ICTI (n=25) |  |  | Comparison |
| --- | --- | --- | --- | --- | --- | --- | --- |
|  | Md | Mean | Sd | Md | Mean | Sd |  |
| 1. How easy did you find it to use the brief cognitive task? | 10.00 | 8.36 | 2.90 | 9.00 | 8.76 | 1.61 | $U=300.5$,<br>$p=0.565$ |
| 2. How helpful did you find the brief cognitive task? | 5.00 | 5.05 | 2.98 | 8.00 | 8.04 | 1.77 | $U=113.5$,<br>$p<0.001$ |
| 3. How burdensome did you find the brief cognitive task? | 7.00 | 6.05 | 3.11 | 7.00 | 6.56 | 1.87 | $U=270.5$,<br>$p=0.931$ |
| 4. How distressing did you find the brief cognitive task? | 10.00 | 9.36 | 1.18 | 8.00 | 8.24 | 1.88 | $U=381.0$,<br>$p<0.05$ |
| 5. Overall, how acceptable did you find the brief cognitive task? | 6.50 | 6.50 | 2.79 | 8.00 | 7.92 | 2.33 | $U=189.0$,<br>$p=0.064$ |
| 8. If you were having intrusive memories in the future, how willing would you be to use the brief cognitive task if it was offered to you as something that would help? | 7.50 | 5.86 | 3.66 | 8.00 | 8.08 | 1.98 | $U=180.5$,<br>$p<0.05$ |
| 9. If a colleague or friend was having intrusive memories, how confident would you be in recommending the brief cognitive task to them? | 6.00 | 5.32 | 3.58 | 8.00 | 7.68 | 2.21 | $U=171.0$,<br>$p<0.05$ |
| 10. How much do you feel that this brief cognitive task could be used within NHS Trusts/healthcare organisations | 5.50 | 5.55 | 3.31 | 8.00 | 8.00 | 2.06 | $U=153.5$,<br>$p<0.01$ |

Note. AFQ was completed at week-4 by AC and ICTI participants only. Quantitative AFQ items 1, 2, 5, 8, 9, and 10 were scored on an 11-point integer scale ranging from 0 ("not at all") to 10 ("very") while items 3 & 4 were scored an 11-point integer scale ranging from 0 ("very") to 10 ("not at all") such that, for all items, higher scores indicate more favourable opinions towards the participant's respective task. The appropriate test was selected based on Shapiro-Wilk normality testing (Mann-Whitney U when violated, Welch's t when met). Items 6, 7, 11 and 12 required free-text responses - the responses were analysed and reported below alongside the interview findings.

### 2.3 Ethical Approval

GAINS-02 received approval from Wales REC 4 (22/WA/0277), with minor amendments including adding qualitative interviews at 12- and 24-weeks.

### 2.4 GAINS-02 RCT Procedures

Recruitment, screening, consent, baseline diary, randomisation, and detailed task procedures are described elsewhere.[15] Participants with work-related trauma and a least 3 intrusive memories per week were randomised to ICTI/AC/TAU. ICTI and AC participants received a single remote researcher-guided session, then had 24-weeks of access of repeatable self-directed task use.

#### Credibility and Expectancy Questionnaire

The CEQ (Table 1) was completed after randomisation and immediately prior to the researcher-guided session for ICTI/AC arms.

#### Task Usage Data

Task-usage data were recorded automatically by the i-spero® system for ICTI and AC across the 24-week period, including number of times tasks were used, duration of task-use, and which intrusive memories were listed and targeted (ICTI only).

#### Acceptability Feedback Questionnaire (AFQ)

ICTI and AC arm participants completed the AFQ four-weeks after the initial researcher-guided session. The AFQ (Table 2) comprises 12 items; eight ratings scored 0 (“not at all”) to 10 (“very”), with burden and distress reverse scored, plus four free-text items (useful aspects, difficulties, suggestions for improvement, and other). Free-text data were analysed by the qualitative researcher (PP) after completion of the interview analysis and were mapped onto the interview-derived themes/subthemes. Responses not fitting existing themes were discussed in relation to whether new themes/subthemes were warranted. Vague or general comments were excluded where theme allocation was not possible. Greater overall acceptability scores for ICTI than AC were reported.[15] Here, we report individual AFQ items for granularity.

#### Qualitative Interview Procedure and Recruitment

When consenting to the main RCT, participants could opt-in to be contacted for a qualitative interview. The interviewer (PP, MSc, Research Associate, female, 6 years of qualitative research experience) was not otherwise involved in the running of the trial, participants had no prior knowledge of or contact with the interviewer, and the interviewer was described to participants as a researcher from the University of Nottingham. PP contacted eligible participants to arrange an interview via audio or video-call. As interviews about usual care were already explored in the GAINS-01 RCT,[13] interviews in GAINS-02 were confined to ICTI and AC. Interviews took place at four-weeks (within a +21-day window post-initial guided session), and between 12- and 24-weeks post-initial guided session (Supplementary Figure S1). After contacting the first 10- to 15 participants, PP used purposive sampling to identify and contact participants from diverse backgrounds (age, gender, ethnicity, job role, location) and with diverse intervention usage (whether participants continued to use the task or log intrusive memories past four-weeks). For the 12-to-24-week sample, participants who did and did not continue using their respective tasks were included to further explore the reasons behind the (lack of) ongoing task usage. There was rapid uptake for interview, so at four-weeks, the first 15 of the 33 contacted participants were interviewed, and at 12–24-weeks, the first 12 participants were interviewed (of 29 contacted) prior to reaching data saturation. Interviews were conducted only once with each participant and participants were not shown their transcripts.

The semi-structured interview guide used at four-weeks (Supplementary Materials) explored participants’ experiences of the study and allocated task. The 12-to-24-week topic guide (Supplementary Materials) explored longer-term intervention usage, such as whether and why participants stopped using the task (e.g. due to no longer experiencing intrusive memories), whether any new traumatic events had occurred, intentions for future use, and implementation for colleagues in the NHS. Interviews lasted between 16- and 51-minutes, and were transcribed and analysed by PP using NVivo14 software.[19] Data were analysed using reflexive thematic analysis[20, 21] with field notes kept throughout and themes generated both inductively and deductively, building on GAINS-01.[13] Longer-term usage and implementation-focused aspects were treated as exploratory given the novel 24-week follow-up in GAINS-02. Analysis involved familiarisation, coding, development and refinement of a coding framework, and iterative team discussion with ongoing reflexive consideration of data interpretation.

## 3. Results

### 3.1 Participants

Between 8 Dec 2022 and 15 Sept 2023, 122 participants were enrolled into the GAINS-02 trial, of which 99 participants were randomised to a treatment arm. Characteristics of the 99 randomised participants are detailed elsewhere. [15] In summary, the main trial sample had a mean age of 41.2 years, were primarily women (86%), and identified as White (90%), with the largest staff group being nursing (46%). The interview sample (N=25) had a mean age 40.6 years, was 92% female and 64% in nursing. Due to selective sampling, the interview sample was more ethnically diverse than the main trial sample (80% White), Supplementary Table S1.

### 3.2 Baseline Credibility and Expectancy of ICTI and AC

As previously reported, there was greater credibility for AC than ICTI - that is, participants in AC were more likely to think their treatment would be helpful than ICTI participants with overall CEQ scores indicating strong evidence of more favourable expectancy ratings for AC (Mean(SD)=35.9 (8.2)) than ICTI (Mean(SD)=32.1 (11.4); Bayes Factor - BF=20.1).[15] Here, we examined individual CEQ items (Table 1) with frequentist statistics. Sub-items showed consistently higher credibility and expectancy for AC than ICTI (numerically), with AC rated as significantly more logical than ICTI (*U*=768.0, *p*<0.05). Mean ratings in both arms were around the mid-range of the scale, indicating both tasks were expected to be only mildly to moderately effective in reducing intrusive memories.

### 3.3 Acceptability of ICTI and AC at 4-weeks

Participants in both arms rated their task as very easy to use (Table 2). ICTI received higher ratings for helpfulness, willingness to use again, confidence recommending to others, and suitability for use within NHS/healthcare organisations. While both tasks were rated feasible and straightforward to use, ICTI was perceived as more beneficial and more suitable for repeated and wider real-world use.

Both tasks were rated as low in distress (higher scores indicating less distress), with AC rated as significantly less distressing than ICTI.

### 3.4 Usage of ICTI and AC over 24-weeks

As reported previously,[15] adherence to the initial researcher-guided session was high in ICTI (97%) and AC (97%), and the mean usage over 24-weeks was comparable between groups. Closer inspection of median usage revealed ICTI was used more frequently in the first four-weeks than AC (Table 3), with median use of 5.0 versus 2.0 sessions including the initial researcher-guided session and a significant between-group difference (*U*=349.5, *p*<0.01). After the first four-weeks, there was little use of either intervention. Median 24-week usage remained higher for ICTI (6.0 sessions) than AC (2.0 sessions; *U*=414.0, *p*<.05). In both arms, overall participant compliance with instructions to engage with the tasks for 20-minutes was high (Table 3).

**Table 3.** Usage data for ICTI and AC conditions.

|  | Active Control (n=35) |  |  | ICTI (n=34) |  |  | Comparison |
| --- | --- | --- | --- | --- | --- | --- | --- |
|  | Median | Mean | SD | Median | Mean | SD |  |
| Number of times either intervention task used |  |  |  |  |  |  |  |
| Weeks 1-4<br>(including initial researcher-guided session) | 2.00 | 2.83 | 2.67 | 5.00 | 6.35 | 6.66 | $U=349.5$,<br>$p<0.01$ |
| Weeks 5-12 | 0.00 | 1.60 | 3.93 | 0.00 | 0.53 | 1.21 | $U=623.0$,<br>$p=0.662$ |
| Weeks 13-24 | 0.00 | 1.14 | 4.43 | 0.00 | 0.03 | 0.17 | $U=664.0$,<br>$p=0.093$ |
| Overall<br>(including initial researcher-guided session) | 2.00 | 5.57 | 9.55 | 6.00 | 6.91 | 7.40 | $U=414.0$,<br>$p<0.05$ |
| Duration of engagement in gameplay or music-listening task in minutes |  |  |  |  |  |  |  |
| Initial researcher-guided session | 20.00 | 19.31 | 1.95 | 20.27 | 20.00 | 2.28 | $U=62.0$,<br>$p<0.001$ |
| Self-guided sessions (AC n=21, ICTI n=26) | 18.46 | 16.16 | 5.24 | 20.59 | 20.28 | 2.94 | $U=57.0$,<br>$p<0.001$ |
| Overall per participant | 20.00 | 17.63 | 3.62 | 20.42 | 20.00 | 3.01 | $U=159.0$,<br>$p<0.001$ |
| Overall per session (AC n=195, ICTI n=235) | 20.00 | 15.12 | 8.26 | 20.35 | 20.78 | 4.02 | ***U=2765.0, p<0.001*** |

Note. The gameplay component of ICTI (i.e., the duration of time for which participants played the game Tetris® using mental rotation) was captured in minutes and seconds. While there were instructions to play for 20-minutes, there was no time limit imposed. The music-listening component of AC could not exceed the duration of the piece (20-minutes). Duration of music-listening was captured in whole integer minutes. Note Tetris® gameplay duration constitutes only one of the five criteria of adherence to ICTI (as outlined in [15], supplementary methods 1) and reported here for comparability to AC task. The appropriate test was selected based on Shapiro-Wilk normality testing (Mann-Whitney U when violated, Welch's t when met).

#### Number of different intrusive memories

On Day 0 (Supplementary Figure S1), ICTI participants listed a median of 4.0 (IQR=3.0-6.00) unique intrusive memories (Table 4). Over 24-weeks, they used ICTI to target a median of 3.00 (IQR=1.00, 4.00), representing 68% (IQR=33%-96%) of intrusive memories initially listed. Participants added a mean of approximately 0.6 new intrusive memories over the 24-week period (median (IQR) is 0.0 (0.0, 0.0)). Most participants therefore did not add intrusive memories after their initial session.

**Table 4.** Summary of intervention use relating to unique intrusive memories, and research contact (ICTI).

|  | ICTI (n=34) |  |  |
| --- | --- | --- | --- |
|  | Md (IQR) | Mean | Sd |
| Number of unique memories listed during initial researcher-guided session | 4.00 (3.00-6.00) | 4.71 | 2.43 |
| Total number of unique memories listed over 24-weeks (including memories added to list independently after initial researcher-guided session) | 4.50 (4.00-6.75) | 5.32 | 2.81 |
| Number of unique intrusive memories targeted with the intervention over 24-weeks | 3.00 (1.00, 4.00) | 3.32 | 2.28 |
| Percentage of all intrusive memories from the intrusive memory list which were targeted with the intervention over the 24-weeks, % | 67.95 (33.33-96.43) | 63.99 | 29.37 |
| Number of times intervention completed per targeted intrusive memory over the 24-weeks | 1.00 (1.00-2.00) | 2.08 | 2.31 |
|  | N | Mean | Sd |
| **Frequency of Researcher Contact to Support ICTI-Use Provided After Researcher Guided Session per participant** |  |  |  |
| Full researcher-guided booster session | n=5 | 0.15 | 0.36 |
| No full researcher-guided booster session | n=29 |  |  |
| Telephone support |  | 0.68 | 0.73 |
| *0 times* | n=16 |  |  |
| *1 time* | n=13 |  |  |
| *2 times* | n=5 |  |  |
| >2 times | n=0 |  |  |

### 3.5 Characteristics of interviewed participants

Supplementary Table S1. presents demographics of the interview sample at 4-weeks and 12-to-24-week timepoints.

### 3.6 Thematic structure derived from the interview data

Five themes were derived from the interviews. Quotes are labelled with participant number (’4_’ = 4-week; ’12_’ = 12–24 weeks) and group (ICTI or AC). Quotes from AFQ are labelled “AFQ”. Additional illustrative quotes are provided in Supplementary Materials. Quotations are provided; quotations in Table 5 demonstrate alignment with the GAINS-01 findings.

**Table 5.** Quotes Replicating Findings from the GAINS-01 Qualitative Study.

| **Theme** | **Quote** |
| --- | --- |
| *Theme 1 – Initial perception of whether to try ICTI* | **Initial scepticism when participants first heard about ICTI**<br>"I was like, "Tetris® are you for real?"... I thought to myself, you know, playing this game's not really going to help me get over this traumatic event. So, I was very, very sceptical in the beginning." (12_010, ICTI) |
| *Theme 2 - Acceptability of ICTI* | "[The intervention] wasn't intrusive, it wasn't something that felt like I had to go out of my way and do and was difficult. It was nice to have it on my phone so that I could just sit and do it on my phone." (4_004, ICTI)<br>"When you sometimes do talk therapies... it's quite emotionally exhausting to go back and talk about them or relive them [the traumatic memories]... that would be a big advantage I think of if it [ICTI] can help address these things and sort of, I don't know re-wire your brain and head in a less exhausting way." (4_002, ICTI)<br>"I did find as the levels obviously got a bit harder and a bit faster, that became a bit stressful and it kind of took away from the relaxing element of playing the game." (4_014, ICTI)<br>"It was easier to fit in the nine to five shift. But then you're trying to fit your life in on those as well because 12-hour shifts leave barely little time for life... It wouldn't be possible [to use at work]. I've barely got 20 minutes to sit down and have anything to eat [on] my night shift just now for example." (4_013, AC) |
| *Theme 3 - Impact of ICTI on intrusive memories and related symptoms* | "As you go to sleep sometimes you think about things don't you, and you'll think about how your day's gone. Now, because I'm leaving it at the door, whereas before I might stay up till midnight, I can actually go to bed earlier. And I'm getting more quality sleep than I did before." (4_010, ICTI)<br>"I think they're less now than they were at the start. And when I get them they're more flashes than staying, if that makes sense... And then you still see [intrusive memory content] but you don't see it for the length of time, so they're becoming shorter and less." (4_010, ICTI) |
| *Theme 4 - Longer-term impact of ICTI on intrusive memories* | "Not saying I won't get anything else in my life, in my work, that might not pop in, but I think it's probably given me a strategy to deal with it, if that makes sense. And it's just knowing that there's something that you can use, and who thought Tetris® would be the thing." (4_010, ICTI) |
| *Theme 5 - Implementation - How to best provide NHS staff with ICTI.* | **Preferences about whether to access ICTI via workplace**<br>"If you are requiring a significant amount of time then really the employer is dutybound, I feel, to allow you that time... I mean if it's a treatment it should be treated as a healthcare appointment, at the end of the day." (4_001, AC) |

#### Theme 1 – Initial perception of whether to try ICTI

##### Lack of awareness of intrusive memories after trauma (the target symptom) and importance of raising better understanding

Participants suggested that 30–100% of NHS staff may experience intrusive memories after psychological trauma, noting this could be an underestimate as many may not recognise this post-trauma symptom. This highlighted the importance of increasing awareness of intrusive memories and of ICTI.

> *“I don’t think a lot of people are aware that they’re experiencing intrusive memories, they just take it as part of the job… I think they’d give it a go if it was something that they’d recognised in themselves was a problem.” (12_008, ICTI)*

##### Initial scepticism when participants first heard about ICTI

Consistent with GAINS-01[13] and pre-intervention CEQ scores (Table 1), participants were initially sceptical that a gameplay intervention could reduce intrusive memories (Table 5).

#### Theme 2 - Acceptability of ICTI

Despite initial scepticism and perceived low credibility before the intervention, participants reported many positives once they began using ICTI. ICTI was described as enjoyable, easy-to-use, and easy to incorporate into their day (Table 5), consistent with GAINS-01.[13] In contrast, despite greater pre-intervention expectancy, AC was perceived as less acceptable than ICTI at four-weeks, particularly as participants recognised it did not affect their intrusive memories.

> *“It was long, time consuming, boring. It had no connection to my intrusive memories. I found no benefit.” (AFQ0072, AC)*

Participants found neither task distressing (Table 2). A perceived advantage of ICTI over alternatives such as talking therapies was not needing to discuss the traumatic memory in detail, instead only bringing the intrusive memory to mind briefly (Table 5).

Participants reported some negatives including difficulty staying engaged for 20-minutes, due to ICTI (Table 5) or AC being irritating, stressful or unenjoyable; However, these issues did not detract from overall acceptability, echoing GAINS-01. Only one ICTI participant reported difficulty focusing on the Tetris® pieces and planning ahead – key components of “mental rotation” when playing Tetris®.

The variety and customisability of the task components were considered important, with participants in each condition suggesting that different games or pieces of music could be used.

> *“I suppose just making it Tetris® might be a bit limiting. Maybe some people prefer a different type of game.” (12_010, ICTI)*

Some people felt they could have done the intervention without human support, some felt optional support would be helpful, and others felt guidance was essential. Support was described as helpful both for recognising intrusive memories and for understanding how to apply the intervention, particularly when breaking complex memories into discrete images. Most participants (10/15) felt they could do the intervention without human support and that optional guidance would be sufficient, supported by existing video tutorials and FAQs, alongside access to email or chat-based support if needed.

> *“Think you could quite have easily put that initial session into either like a short interactive tutorial or a video… I think having that reassurance of some optional assistance probably best.” (4_005, ICTI)*
>
> *“I think you have to have it guided because I really struggled with it… I wouldn’t have understood what they were trying to get at saying, “You have to break them [memories] down into single sort of images.” I was just like, “Well they’re not images they’re running memories, they’re like movie scenes and you get caught up in the movie scene all the time.”” (4_004, ICTI)*

Some participants reported difficulty finding 20 minutes to use ICTI/AC, due to work and personal responsibilities (Table 5).

#### Theme 3 - Impact of ICTI on intrusive memories and related symptoms

Participants from both groups noted benefits from using their respective task on improved sleep and focus. Data reported in the GAINS-02 RCT [15] showed ’strong’ evidence of lower insomnia symptoms at weeks-12 and 24 after ICTI relative to AC. Some AC participants reported the task helped them relax or functioned as distraction, while ICTI participants more typically reported benefits related to no longer experiencing intrusive memories, including reduced avoidance of certain situations (Table 5).

> *“Yes. I’m not feeling like I need to avoid certain scenarios, and not stressing, is something going to pop into my head? I just haven’t got that worry now about the stuff behind the memories or the memories coming back.” (12_003, ICTI)*

ICTI participants reported their task helping with the target symptom (Table 5; a reduction in frequency or intensity of intrusive memories), convergent with the main trial results.[15] Many AC participants recognised that their task, while potentially relaxing, did not target the key symptom. This aligns with the main trial data showing substantially greater reductions in intrusive memories in the ICTI arm than the AC arm over 24-weeks, with 70% of ICTI participants symptom free at follow-up compared to 13% in AC.

> *“I think it is a relaxing, that helps you sort of be more mindful and that’s a big help with that. But I don’t know whether it was directly linked to intrusive thoughts or whether it was just nice to use it.” (12_006, AC)*

#### Theme 4 - Longer-term impact of ICTI on intrusive memories

Many ICTI participants did not need to continue the intervention beyond the first four-weeks, as they were no longer experiencing intrusive memories (Table 3). In contrast, reduced usage by AC participants reflected finding the task unhelpful or forgetting to use it.

> *“I haven’t needed to [continue use] because I haven’t had any memories that were troubling or anything.” (12_008, ICTI)*
>
> *“On and off. But I think it’s quite hard, like, with work getting in the way and just, like, reminding myself to.” (12_012, AC)*

ICTI participants highlighted they would use the task again in the future should new intrusive memories appear, having seen that it was effective (Table 5). Whereas AC participants willing to use the task again did not cite help with intrusive memories as the reason.

#### Theme 5 - Implementation - How to best provide NHS staff with ICTI

##### Preferences about whether to access ICTI via workplace

Given that ICTI was found to be highly successful in reducing intrusive memories[15] and acceptable, we explored future implementation. Participants described advantages of workplace delivery (Table 5), including being allowed time at work to use interventions, as previously reported in GAINS-01.[13] Additional advantages included employer funding and perceived legal or occupational protection.

> *“I think it has to come through work because for the very nature of you know what I’m still being asked to do… Did I do everything right? Going to the coroner. The scrutiny of notes… Will I still have an income? So, there’s a whole trail of negativity associated with very difficult stressful events at work. It doesn’t just end with what you’re seeing or witnessing, it’s your own self-preservation and your livelihood.” (4_001, AC)*

In GAINS-01, mental health stigma was identified as a key barrier to accessing workplace interventions (Table 5). This was echoed in GAINS-02, although some participants felt stigma was reducing.

> *“You feel like you’re a failure if you’re not on top of it and you’re not coping and that kind of thing. And that’s awful, isn’t it? But actually, it’s probably in an indirect way from COVID and everything. It’s made everybody able to talk about it and how awful it was, and you know so.*
>
> *Normalised it a little bit more in that way, so yeah. It’s an acceptable thing to be stressed about.” (12_006, AC)*

Despite perceived reductions in stigma, some participants preferred to access interventions outside the workplace, or via routes where their identity would not be known. Reasons included difficulty accessing or trusting Occupational Health and discomfort discussing symptoms with line managers.

> *“I don’t really want work to know. It’s terrible because I mean obviously, I work for the healthcare profession, you know, looking after your workers is part of it. But yes, I don’t know, I just feel that if it was on my permanent record that it might, I mean it’s not supposed to, but it might like be detrimental in the future.” (12_002, ICTI)*

When asked about access to their data, most participants preferred access outside the workplace (e.g. via their GP), with only one identifying Occupational Health as helpful.

> *“I’m not sure I’d want work to have access to that all. I don’t know that they’d need to… I guess if somebody had to, I think I’d rather it be my GP.” (4_014, ICTI)*

Participants emphasised that preferences for workplace delivery compared to access outside the workplace would vary by individual and organisational context.

> *“So, I think if you offered it through both ways [within and outside of the workplace] and then it’s universal, isn’t it? So, if you’re a bit embarrassed about talking about the Occupational Health about it, you can come away with that code and just scan it and you can access it without anybody knowing basically.” (4_015, ICTI)*

##### How to best get NHS management on-board with ICTI

Participants suggested that implementation should be pitched to NHS decision-makers in terms of improved retention and reduced sickness absence from an intervention that requires minimal resourcing.

> *“When it first happened, I took a lot of time off to get over it and get my head straight, which costs a lot of money to the NHS. I think when you are approaching people like that, you need to go at it from a business point of view of saving money, preventing occupational health referrals, these sorts of things. Staff not leaving, so staff retention.” (12_008, ICTI)*

Participants suggested that national-level approval (e.g. NHS England), rather than Trust-by-Trust adoption, would facilitate implementation, alongside targeting teams responsible for staff wellbeing.

> *“You don’t need each individual Trust to take it up you just need NHS England to take it up. So once they say yes all the Trusts have to do it. It’s like the wellbeing champions the hospital have a very big commitment to it because the NHS do and the NHS have told them to.” (12_004, AC)*
>
> *“Maybe if you train more people in house, specifically… say like mental health first aiders, I think some type of help might be all it needs.” (12_006, AC)*

##### Advantages of ICTI over existing NHS wellbeing provision

Participants reported largely negative experiences with existing workplace wellbeing initiatives, describing them as poorly timed, poorly located, or irrelevant. ICTI was perceived as more relevant and accessible.

> *“So many of these other well-being incentives, especially in the hospitals, come from managers who just seem to have no concept of what it is actually like. Although I don’t know who actually developed this [intervention], but this feels so much easier and it sounds ridiculous but relatable… it feels like someone has actually thought about the science behind stuff” (12_003, ICTI)*

## 4. Discussion

Despite initial scepticism and weak expectations about the likely effectiveness of both ICTI and AC, five uses of ICTI (approximately once per unique intrusive memory listed) in the 24-weeks after the initial researcher-guided session was usually sufficient to stop intrusive memories recurring. ICTI participants reported that reductions in intrusive memories carried further benefits, including improved sleep quality and reduced avoidance of trauma-related places, aligning with quantitative findings.[15] In contrast, participants found that AC had little direct effect on traumatic memories although it did serve as a distraction. Participants did not use AC much during the six months because they found it ineffective. Participants described ICTI as a strategy they could reuse for new intrusive memories, consistent with prior work suggesting value for staff exposed to ongoing trauma.[11, 12]

Despite their initial scepticism, once participants tried ICTI, experienced benefits translated into greater willingness to reuse the intervention and recommend it to colleagues and NHS organisation, with a felt sense that ’it’s *weird but it works*’ (JH). Consistent with prior work,[13] participants emphasized advantages over existing treatments, including being less ’*emotionally exhausting’* than psychological therapies, due to not needing to talk about the traumatic memory. Additionally, ICTI requires less time than typical psychological interventions (e.g. 12-20 hours[22]). This is of interest given that 70% of ICTI participants reported no intrusive memories by 24-weeks after a median of six sessions. While guided support sessions and short video instructions were valued, many participants felt they could use it without a guide, and optional guided support would suffice, strengthening the value proposition for time-pressured staff and supporting scalability. Overall, this low-distress profile of ICTI, brevity and ease of use within a digital platform may support engagement and broader implementation.

When considering implementation, some participants felt interventions to support staff mental health should be available through the workplace as part of employers’ duty of care, given intrusive memories are an occupational hazard. Intrusive memories cause harm even in the absence of diagnosable disorders such as PTSD, with staff highlighting limited awareness about intrusive memories of trauma, indicating a future need for education about their nature and impact. While some participants were open to accessing ICTI via their employer, others preferred multiple access routes due to concerns about privacy, anonymity, and trust in the employer, consistent with findings from other workplace interventions.[13, 23]

By interviewing both continued users and non-users and triangulating interviews with acceptability, expectancy and usage data, we gained a more detailed picture of how ICTI was used and how uptake might be improved. Integration of the qualitative and quantitative data occurred only at the interpretative stage of the project, so it is unlikely that integration of the data in this way biased responses. Replications across UK[13] and Sweden[14] suggest robust effects and similar implementation challenges across healthcare systems. A key limitation is that we could not interview eligible staff who declined participation in the study. This group warrants further research, particularly as some staff may not recognise intrusive memories nor understand the rationale or need for intervention. Furthermore, due to the pronounced attrition at week 4 across all measures,[15] we have AFQ data for just 47 out of the 69 participants. Those who did not complete the AFQ may have had a different, potentially less positive, experience to that which is reported in this paper.

Future work should evaluate cost-effectiveness of ICTI, and more closely examine whether reductions in intrusive memories translate into improvements in burnout, sickness absence and staff retention. An implementation study is needed to estimate real-world uptake, identify barriers and facilitators, and develop programme theory clarifying which contextual factors shape outcomes. This should include testing a self-guided model, given that many interview participants felt optional guided support would be sufficient. A self-guided approach, incorporating participant recommendations such as video demonstrations of someone identifying the intrusive memory images independently and playing the intervention while focusing on mental rotation,[13] could enable greater scalability of this intervention.

## Conclusion

ICTI was perceived as highly acceptable and effective after psychological trauma, with ICTI participants frequently reporting task-specific reductions in the frequency or intensity of intrusive memories in their interviews, consistent with main trial findings[15] Perceived advantages included ease of use, brevity, requiring minimal guidance, the non-distressing nature of the intervention, and strong potential for scalable implementation among healthcare staff. Findings were triangulated after separate analyses to add contextual richness to the quantitative findings to inform implementation work on ICTI, scalability and potential adoption into routine care. Future work should assess cost effectiveness, impacts on presenteeism and retention, and real-world implementation including the feasibility of a self-guided ICTI.

## Contributors

PP carried out the interviews, collected and formally analysed the data, and drafted, reviewed and edited the paper as first author.

RM, EAH, AM AB, MF and SB contributed to methodology, writing, review, and editing. MF contributed to formal analysis of the quantitative data.

EAH and JK contributed to conceptualization, funding acquisition, project administration, methodology, writing, review, and editing.

CS contributed to conceptualization, review, and editing. JH contributed to review and editing.

## Data and materials availability

The raw interview data collected in the interviews cannot be made available in an open access format. Participants did not provide consent for their full transcripts or recordings to be shared publicly, and many of the narratives include personal details that could make individuals identifiable even after standard anonymisation procedures. To protect participant confidentiality and uphold our ethical obligations, access to the raw data must therefore remain restricted. Data for the statistical analysis is available at: https://osf.io/cs6hn/files/osfstorage

## Acknowledgments

This work was primarily supported by the Wellcome Trust (223016/Z/21/Z). The funder of the study had no role in the conceptualization, design, data collection, analysis, decision to publish, or preparation of the manuscript. EAH also received funding from the Swedish Research Council (2020–00873). CS salary was partly funded by National Institute for Health Research (NIHR133788) and UK Research and Innovation-Medical Research Council (MR/S035753/1 and MR/X005070/1). PP was partly funded by NIHR Applied Research Collaboration East Midlands (ARC EM). RM was partly funded by NIHR ARC EM, the Nottingham NIHR Biomedical Research Centre, the NIHR Mental Health (MindTech) HealthTech Research Centre and a NIHR Senior Investigator Award. The authors thank all participants for their role in the study. The authors thank the Intensive Care Society; our Data Monitoring Committee members including Andreas Reif (chair, Frankfurt am Main—Goethe University), Steve Hollon (Vanderbilt University), and Ian Penton-Voak (Bristol University); our Trial Steering Committee; Expert Advisors including Nick Grey, Simon Wessely, and Jonathon Bisson; and all members of the study team at P1vital Products Ltd. We are grateful to The Tetris Company, Inc for their support in licensing the Tetris^®^ game for use within i-spero^®^ as a component of digital ICTI in this research.

## AI Use Disclosure Statement

Artificial intelligence tools were used only to support minor editorial tasks during manuscript preparation. Specifically, AI was employed to suggest areas where word count could be reduced and to assist in formatting and producing references. No AI tools were used for data analysis, interpretation or drafting of substantive content. All final decisions were made by the authors, following review of changes suggested by AI.

