## Supplementary Materials for "Trauma in healthcare staff: A multiple methods study using quantitative and qualitative lived experience of participants in a randomised controlled trial of a brief digital imagery-competing task intervention for intrusive memories"

### Figure S1. Flowchart of GAINS-02 Study Procedures

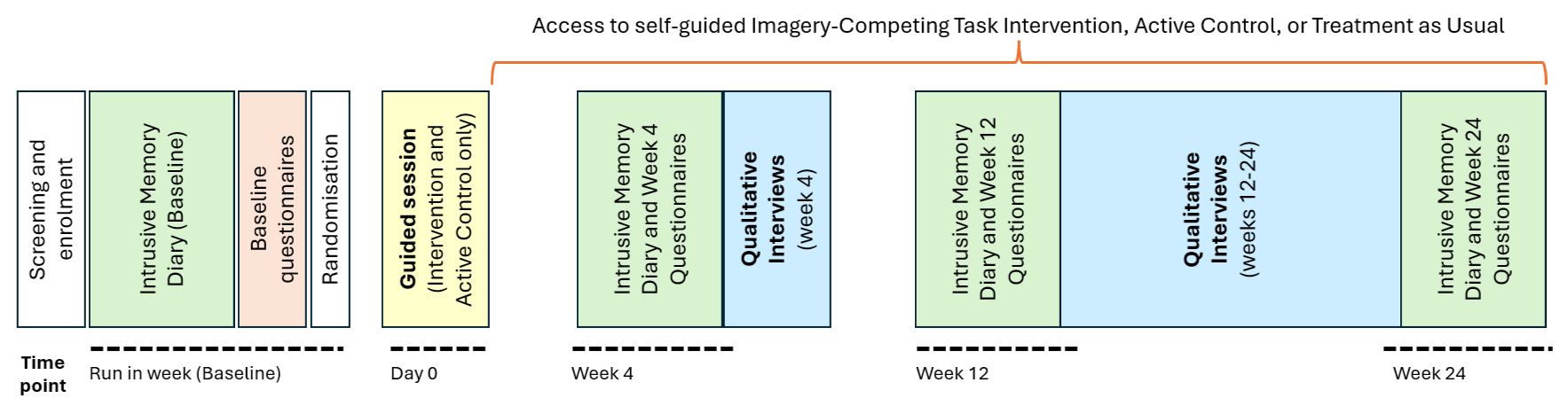

**Figure 1. Flowchart of GAINS-02 study procedures to indicate current data collection.**

**Note,** *credibility and expectancy questionnaire for all study arms was completed after randomisation on Day 0 (and for the two intervention arms this was immediately prior to receiving allocated intervention in the researcher-guided session); the acceptability feedback questionnaire was completed at week 4 (ICTI and active control arms only): task-usage data were collected during the initial guided session and over the 24-week in-study period (ICTI and active control arms only). Qualitative interviews were conducted after 4-weeks and between 12 to 24-weeks (ICTI and active control arms only).*

### Table S1. Demographics of Interview Sample.

Data presented for N=25 participants in total, with 27 interviews.

|  | **4-week sample**  **(n=15)** | **12- to 24-week sample**  **(n=12)** | **Total**  **(n=25^1^)** |
| --- | --- | --- | --- |
| **Arm** |  |  |  |
| ICTI | 8 | 8 | 14 |
| AC | 7 | 4 | 11 |
| **Age** |  |  |  |
| 18-25 | 0 | 2 | 2 |
| 26-35 | 5 | 4 | 8 |
| 36-45 | 3 | 2 | 5 |
| 46-55 | 6 | 2 | 7 |
| 56+ | 1 | 2 | 3 |
| Mean | 42.3 | 38.5 | 40.6 |
| SD | 10.1 | 13.6 | 11.6 |
| **Gender** |  |  |  |
| Male | 2 | 0 | 2 |
| Female | 13 | 12 | 23 |
| Other | 0 | 0 | 0 |
| **Ethnicity** |  |  |  |
| White | 12 | 10 | 20 |
| Mixed | 1 | 1 | 2 |
| Chinese | 0 | 1 | 1 |
| Other | 1 | 0 | 1 |
| Asian | 1 | 0 | 1 |
| **Job Role** |  |  |  |
| Allied Health Professional | 3 | 1 | 4 |
| Doctor | 1 | 0 | 1 |
| Health informatics | 0 | 0 | 0 |
| Healthcare Support Worker | 0 | 2 | 2 |
| Nursing | 10 | 8 | 16 |
| Other/unknown | 0 | 1 | 1 |
| Pharmacy | 0 | 0 | 0 |
| Missing | 1 | 0 | 1 |
| **Number of intrusive memories in Baseline week** |  |  |  |
| 3-5 | 2 | 3 | 4 |
| 6-10 | 5 | 3 | 8 |
| 11-15 | 3 | 1 | 4 |
| 16-20 | 2 | 3 | 5 |
| 21-25 | 1 | 0 | 1 |
| 26-30 | 1 | 1 | 1 |
| 31-35 | 1 | 1 | 2 |
| **Number of intrusive memories in Week 4** |  |  |  |
| 0-5 | 11 | 10 | 20 |
| 6-10 | 2 | 1 | 2 |
| 11-15 | 2 | 0 | 2 |
| 16-20 | 0 | 1 | 1 |

Note. **^1^***The total number of unique participants is 25 rather than 27, as 2 participants completed both the 4-week and 12- to 24-week interview, so they are only counted here once.*

### Table S2. Additional Quotes Illustrating Themes Identified from Analysis

| **Theme** | **Quotes** |
| --- | --- |
| Theme 1 – Initial perception of whether to try the ICTI | “Yes, I think I was surprised a little because obviously I wasn’t initially even sure that I’d be eligible for the study or it was even really a thing that I was having, and then when, I think, one of your colleagues, sort of, you know, you do the education to go through that and then when we started actually breaking it down I think I realised that I had more images coming up than I ever thought… I think there’s probably a bunch of people out there like me who don’t even realise that they’re having some issues, probably, I don’t know.” (4_002, ICTI)  “I suppose I was probably a bit skeptical in a similar way as I’d probably been skeptical about lots of things that have happened over the last few years in terms of, you know, when I first started talking therapy, because I didn’t think it would work, but it did.  And EMDR I thought, I’d obviously heard about it before, I just thought it all quite bizarre, but again, I went into with an open mind and that definitely helped.” (4_005, ICTI)  “Yes. So I was very sceptical in the beginning and the other research introduced this Tetris and I was like, “Tetris are you for real?” Because I didn’t really think of it as- So I’ve have PTSD from another thing years ago so I thought to myself, you know, playing this game’s not really going to help me get over this traumatic event. So I was very, very sceptical in the beginning.” (12_010, ICTI)  “When I told my husband I was taking part in a research project regarding memories and I’ve got to play Tetris. Even he was a bit like, you’ve got to play Tetris? Like that.” (4_007, ICTI)  “I would say at least 60% of the people that I’ve heard like actually we’re looking at more like 75% of people I’ve worked with. They haven’t necessarily said to me, “This is what happened and this is how I feel about it,” but we talk about random things even on our lunch breaks we’ll talk about and we’ll joke about it sometimes, but we all recognise deep down that it’s quite traumatic.” (12_010, ICTI)  “They probably wouldn’t bring it up as intrusive memories or PTSD or anything along those lines, but you could ask them and they will say I get a little feeling when I go onto ICU or when I go onto [ward name] there was something that happened and there was always one thing that affected you.” (12_004, AC)  “When I told my husband I was taking part in a research project regarding memories and I’ve got to play Tetris. Even he was a bit like, you’ve got to play Tetris?” (4_007, ICTI) |
| Theme 2 – Acceptability of the ICTI | “It was fun, it didn't feel like I was working on something distressing or triggering but it still worked.” (IFQ0113, ICTI)  “I had a memory pop up in the middle of the day, so I opened the game and played there and then. Super easy to do and it allowed me to refocus.” (IFQ071, ICTI)  “In the very beginning I was like really struggling to mentally find the time I didn’t see it as something important because obviously I hadn’t seen the efficacy of it. But as I experienced it and it was positive I thought, Okay well this is great. Yes, I’m just going to do my little 20 minutes of therapy for the day. It felt like that was the push factor for me to do it.” (12_010, ICTI)  “I think the thing that I found trickier, but again when I was guided by one of your colleagues it was easier was just breaking those images down- Not that he actually told me what to write but then in the way to do that and separate them out. So it was probably more just that assistance with the breaking down the images… but I suppose that was, maybe, if anything, a harder aspect, working those bits out, the initial images to tackle.” (4_002, ICTI)  “It needs to be far more brief. This is aimed at healthcare staff who are trying to deal with intrusive memories while working with patients or other busy settings. They can't just stop for 20 minutes every time they get a distressing thought.” (IFQ057, AC)  “Therapy is scary it’s another human it’s structured it’s not you it’s not just you escaping.” (12_004, AC)  “I suppose just the fact that you know they might not want to go on medication. So, it’s an alternative healing isn’t it?” (12_005, ICTI)  “Tetris is easy, I’ve been playing that since I was a kid so that was great [laughs]. Once you started it went really quickly and it was enjoyable actually and fine.” (4_002, ICTI)  “I think I’d just be realistic that it isn’t just playing a game. There is a little bit of it that’s like, “Oh, I’m going to have to think about it,” but it’s worth it… I was never too distressed by bringing them into mind.” (12_007, ICTI)  “I think the task was quite frustrating but only because you lose focus around 15 mins mark.” (AFQ0125, ICTI)  “There are other pieces of music I would have enjoyed listening to more, so I don’t know if it did anything for me. It felt a little bit like a task, like “Oh, I’ve got to listen to this and I’ve got to listen to it to the end,” sort of thing.” (4_009, AC)  “Maybe you’ve got similar ones that you can present, so you’re not listening to the same thing all the time. Because music’s quite personal to people, isn’t it. Especially genre types.” (4_001, AC)  “You’ve still got that mental rotation with Bubble Pop because you’re looking at the colours and you’ve got to match it, so it’s a bit like, it is like Tetris, but it’s a bit more fun.” (12_009, ICTI)  “Because I do a lot of knitting and crocheting and things like that, so they said, “Why don’t you try it with knitting and crocheting? Because it’s the same sort of repetitive action”. And I found that quite useful… you’ve got to sort of pre-plan your next sort of stitches if you’re putting them in and where they go.” (4_004, ICTI)  “The issues were just mainly time, getting it done, because I have like three children… by the time I get everyone to bed it’s like 10 o’clock and I’m exhausted. I just go to bed and then I think, “Oh no I haven’t done it”… and then I fall asleep and then it’s like I didn’t actually do the intervention.” (4_004, ICTI) |
| Theme 3 - Impact of the ICTI on intrusive memories and related symptoms | “I think it takes away the urgency to enable you to focus on what it is that’s actually bothering you in order for you to work on it, rather than being in that flight and fight response… and I suppose recognizing that, it gives you a bit of a control.” (4_015, ICTI)  “Yes, I feel like it’s almost a like a cushion that I can fall back on if I needed to now that I have this sort of- It’s a coping mechanism I see it as a coping mechanism now.” (12_010, ICTI)  “I think they’re [intrusive memories] less now than they were at the start. And when I get them they’re more flashes than staying, if that makes sense… And then you still see [intrusive memory content] but you don’t see it for the length of time, so they’re becoming shorter and less.” (4_010, ICTI)  “They’re [intrusive memories] pretty much entirely gone. And like I- I mean, it’s kind of unbelievable that they’ve gone.” (12_007, ICTI)  “I think it takes away the urgency to enable you to focus on what it is that’s actually bothering you in order for you to work on it, rather than being in that flight and fight response… and I suppose recognizing that, it gives you a bit of a control.” (4_015, ICTI)  “Whenever I was working on ICU during Covid I couldn’t sleep at night because everything was just rolling over in my head but from this, I was able to decompress quicker or something, I don’t know.  It felt like when I was listening to the music it was just like taking a big deep breath in and letting it all go.” (4_008, AC)  “It redirects my attention or it shapes my focus from the bad memories to a more positive experience” (4_011, AC)  “There was one that used to come up more often… I haven’t really thought of it or it hasn’t popped in [laughs], I don’t know if that’s because I’ve been really busy but I think probably the intervention has helped.” (4_002, ICTI)  “Initially I was targeting two, but then I was targeting up to four. But now it's gone down to I'm literally targeting one now. It's not daily either. Now it's more so it'll come, but it's not as intrusive, not enough to record it. When it is kind of intense, then I will record it and do the- And use the task as well. They haven't disappeared, they're still there but they're not as prominent and they're definitely not as clear. They wouldn't warrant me to say, that's really bothered me today. I should really report that, if that makes sense.” (4_015, ICTI)  “I had an initial drop [in intrusive memories]... this week possibly a couple due to some traumatic events at work.  But they’re back again.” (4_001, AC)  “I knew it wouldn’t be helpful… I mean in initially I was quite good at-  I was really engaged and mentally good, but I realised quite quickly that the music and the intervention that I’d been given wasn’t going to help at all. It wasn’t going to encapsulate, it wasn’t linked to my intrusive thoughts or anything .” (12_001, AC) |
| Theme 4 - Longer-term impact of the ICTI on intrusive memories | “But yes the intrusive thoughts since I’ve done the intervention- I mean I’ve nipped in just because I quite like Tetris and just like when I’ve had triggers at work I’ve been and done just a couple of sessions on it just to stop any intrusive thoughts coming back again.” (12_002, ICTI)  “They’re pretty much entirely gone. And like I- I mean, it’s kind of unbelievable that they’ve gone.” (12_007, ICTI)  “It’s easy to incorporate into everyday life without having to take time out to do something. I think that would be the main reason.” (12_012, AC)  “So, I mean, since the four weeks I’ve probably only used the intervention twice… since I’ve been asked to record if have any intrusive memories I’ve not had any. So I haven’t really needed to use the intervention for probably the last four to six weeks.” (12_010, ICTI)  “The whole thing wasn’t to link it or bring that thought into my mind or the memory or anything, just to listen to it at any time of the day if I want. And that was the only instruction, so I quickly realised that it’s not going to help me, so I listen to the music, I know what it sounds like, listened to it a few times, and then I just put it out of my head.” (12_001, AC)  “Yes, I feel like it’s almost a like a cushion that I can fall back on if I needed to now that I have this sort of- It’s a coping mechanism I see it as a coping mechanism now.” (12_010, ICTI)  “No. Because it wasn’t linked to my intrusive thoughts or my trauma. There was no correlation between the two so it was not going to be helpful at all. I know the piece of music was long and it would have been very boring to have listened to it every day for no benefit at the end.” (12_001, AC) |
| Theme 5 - How to best provide NHS staff with the ICTI, i.e. implementation | “We’re coming in to an era where this sort of thing is much more in the forefront of people’s minds. There’s an awful lot about mindfulness and you know wellbeing and kindness in the workplace and sort of there’s you know, there’s a lot of work going on and it’s the forefront of the media at the moment.” (4_003, ICTI)  “Occupational health probably feels quite a distance away… Like, our occupational health is on another site so it’s a 20-minute drive away… They have to ring up, they have to make an appointment, they have to drive to it, so they’re not going to do that. They will go off sick before they go to occupational health.” (4_010, ICTI)  “I think it does depend where you work.  And going back to you know where I was, where I experienced all these awful things, I wasn’t necessarily sharing that sort of information there, just wouldn’t dream of it.” (4_001, AC)  “I think it has to come through work because for the very nature of you know what I’m still being asked to do… Did I do everything right?  Going to the coroner.  The scrutiny of notes.  Is my VIN number safe?  Will I still have an income?  So there’s a whole trail of negativity associated with very difficult stressful events at work.  It doesn’t just end with what you’re seeing or witnessing, it’s your own self-preservation and your livelihood.  It’s my default setting -  I think if you talk to any healthcare professionals, we’re guilty until proven innocent.” (4_001, AC)  “I think people think it’s just almost lip service. Rather than actually putting things in that would improve everybody’s wellbeing at work on a day-to-day basis when there are people who are just asking for literally a chair to sit on and they’re like, Have a wellness teabag.  You’ve got doctors on the ward sitting on bins because they have nowhere else to sit.  You just think, This is what you think is going to help people?  So I think it needs to be an external thing.” (4_013, AC)  “But I would personally I would make it separate from work because then you’re never going to read the data privacy sheet and so I think people might think, Well on the 1% chance that my employer might be able to access this information I’m not going to do it. Just in case, you know, we go into this whole world where employers can see stuff and then fire you on that basis.” (12_010, ICTI)  “I wouldn’t feel comfortable raising that with my line manager and being, like, “I am struggling.”… And I think it probably depends, like, on your line manager, the department you work in, and everything like that… And, like, if you’re woman it’s, like, oh, you’re, like, seen as weaker probably… I’m only speaking from my experience.” (12_012, AC)  “Because very often we just feel we are treated like numbers by a lot of senior management so if there’s more of an initiative to make sure that we’re well so that our care is good… Because there’s so much blame culture within the NHS so if they try and take that pressure that, “Absolutely we recognise that you’re burnt out and you’re struggling so we want to do something for you so that you’re in a better place to be a better- Because we know that you’re a good practitioner we know you’re not unsafe we just- Under too much pressure.”” (12_010, ICTI)  “The NHS has so many problems already I feel like unless like everyone like 75% of the work staff has this problem and labels it, I don’t think they would really like do anything about it because there’s so many more like pressing issues.” (12_011, ICTI)  “Most of the time when you read like the wellbeing board and whatever it’s just really limp like the stuff that’s on there... Like you wouldn’t really want to get involved in it unless you’re really struggling... So I think this is a step above what we would have in wellbeing at work things and stuff like that because it’s actually looking at the science about it. And that’s going to appeal to the healthcare professionals… It’s worded in a way about what our brain does and how our brain works I think that people would be a little bit more inclined to take note of it and actually think it’s actually an effective tool. Rather than it just being like, you know, “If you sit down every Wednesday and talk to some random stranger about your personal feelings we’ll enter you into a raffle.”” (12_010, ICTI)  “When it’s through the workplace it’s like free or like subsidised, but when it’s private like people would generally charge.” (12_011, ICTI)  “And then I do wonder about more of a like a real big culture shift and that’s in terms of looking at shift patterns and time allocated actually when the shift finishes half an hour early and there’s time, or like 15 minutes early and there’s time dedicated to doing some sort of intervention… You have to have protected time… it goes right up to policy and like even the protection in law that has been talked about in terms of staff ratios.” (4_002, ICTI)  “There is the stigma around, you know, especially with nurses, you know, we’re supposed to be strong… Because I know one of the girls she was off… she felt so much guilt, you know, because the added pressure of one less member of staff at work, but also felt guilty for going to work because she was leaving her children. She had all this pressure, and I was like, “But you need to take time for you. You need to look after you.” (12_002, ICTI)  “I think [the workplace having access is] quite important. Because occupational health could put a bit of a levy on management, that if the individual was struggling to cope in a particular area then they can be transferred without loss of a grade or income.” (4_001, AC)  “I would have a preference of outside of work, but I know a lot of my colleagues access occupational health inside of work, so I wonder if both could be offered, that there’s two ways of actually accessing it. So like, you could do it yourself, but also do it through recommendations from occupational health.” (12_009, ICTI)  “Decreasing the amount of unnecessary sickness leave then that would be a very good way of demonstrating that it works. I think a lot of people that work in trusts in senior management they’re very sceptical and they can’t- They don’t have- They can’t afford to take a lot of risks really… and there isn’t a lot of funding to go around sometimes especially not for staff wellbeing. There was a question about, “Have you take many days off work?” That could be a really useful- Because I haven’t. I’ve actually been so much better at work like I- I used to call in sick quite a lot before my shift. So I don’t know whether that’s directly related to this intervention and since I’ve been using this intervention I’ve actually wanted to go to work most of the time.” (12_010, ICTI)  “They do not understand the importance of portering and they are ripping the soul out of it. So if you went to them with this idea they would just turn round and laugh at you because why would they want to look after us is the way I feel at the moment. Luckily they are not the people that would be implementing it. The leader of the Mental Health Champion Scheme is called [leader's name] and she would definitely be interested. I have spoken to her about what I’m doing and she finds the idea interesting and they would be the ones that … that have a mental health team that would implement it.” (12_004, AC)  “All these other workshops and stuff that they do, you’ve got to have a full day to do it or you’ve got to come off the floor for two hours to go and do a study thing, whereas this isn’t going to take away from that.  Then people start feeling better and feeling that they can deal with stuff better and they have tools that are easy to use and access, they work better… it’s really hard to get time off for study days and stuff so it doesn’t require that.” (12_003, ICTI) |

### Topic Guides for Semi-Structured Interviews

#### 4-week Interview Topic Guide

**Overall views**

- In this study you used an intervention, which included completing a brief imagery completing task / brief music-listening task. What are your views on this intervention, how did you find it?

**Impact**

- Have you experienced a change in the number or frequency of intrusive memories you are experiencing?
- What are some of the problems caused by intrusive memories for you, and to what extent has the intervention helped?
- Has the intervention helped you in any other ways?
- What is your main motivation for using the intervention (e.g. improvement in your own health and wellbeing, or by improvements in how you are doing at work or ability to deliver quality care, or something else)?
- Have there been any unanticipated changes as a result of using it? Immediate or over time (this could be anything, so best to leave open). If asked for prompts, can offer: from relaxing / fun, to starting to play more games / playing it with family, gaining insight from their logging of memories, emotions being triggered by logging of memories, being put off Tetris as a game, associating phone or Tetris with bad memories….

**Ease of use**

- How easy was it to use the intervention? What was easy or helpful about it?
- Did you experience any difficulties with accessing or using the intervention?
  - Did you dislike or avoid using it for any reason?
  - Do you have thoughts on how we could overcome some of those problems?
- How easy or difficult was it to incorporate this digital imagery competing task or brief music-listening task into your day?

**Type of use**

- How did you use the digital imagery competing task or brief music-listening task?
  - How often did you use it?
  - At what times and under what circumstances did you use the digital imagery competing task or brief music-listening task? When and where did you use it?
  - If not often, why was this?

**Support (first session)**

- In this study a researcher helped you get on to the system and use the intervention for the first time. That will have happened in a phone call or videocall. How did you find that session?
  - Do you think it worked well as a way of getting set up and started with the intervention?
  - How clear were the instructions provided?
  - How could it have been improved?
  - Did you feel confident about using mental rotation appropriately?
  - Did you feel like you understood enough about the intervention itself (i.e. how it works), to understand how it might benefit you?
  - Did you feel you understood enough about the intervention to have faith in it being helpful to you?
  - Did you believe the intervention would bring you benefits that were important to you? Did you have any scepticism about the intervention?
  - Overall, did you feel you had enough support to start using the intervention effectively?

**Support (additional support)**

- After you were set up and have completed the intervention the first time, did you receive any additional researcher support? What were the circumstances of this taking place?
  - If yes, why did you feel the support was required?
  - What did this involve and how helpful was it?
  - If no, did you feel comfortable accessing the intervention on your own without researcher support?

**Support (independent use)**

- How would you feel about using the intervention entirely independently, without assistance?
- What would help you or others to use the intervention independently?
- Did you identify images on your own? How did you feel (if ‘yes’) or would you feel (if ‘no’) about identifying images on your own? Did you require any support with this? What would help you/others to identify images independently or play Tetris in the right way?
- How would you prefer to use the intervention - entirely independently, with assistance or with optional assistance?
- Did you have any problems using the intervention, that we have not already covered?

*(The following questions are optional and only to be asked if earlier parts of the interview have already been addressed)*

- We are interested in cases where people might benefit from the intervention but for whatever reason cannot or choose not to use it. Thinking about the people that you work with, are there any examples of where that might happen?
- Do you think there might be other barriers to colleagues using this intervention? What might these be? E.g. practical (device access / time); cultural (don’t like games, stigma); related to ability (eyesight, hearing, manual dexterity, cognitive/psychological ability); privacy / anonymity (need to access via non-work routes, or be packaged without references to ‘problem’; confidence in safety of data)
  - Are there any groups of colleagues who may be more or less likely to use it, and what might make them more likely?
  - How might we be able to overcome these inequalities in access? (if relevant)

**Study processes**

- Which NHS Trust do you work in? (purely asked for selective sampling purposes)
- Did you find the intervention burdensome / distressing? If so, what aspects specifically?
- How did you find filling in a diary with your intrusive memories every day? Did you find it helpful or unhelpful?
- Is there anything that could help to reduce the burden / distress caused by this intervention?
- How helpful were the intrusive memory graphs? How did you use them?
- What are your thoughts on the reminders? Did you find them helpful? (Why / why not?)

**Stigma / comparison to other treatments**

- Other than this intervention, have you ever sought help for intrusive memories? For example, speaking to your GP, talking therapies, or medication?
  - If yes: How does this digital imagery competing task or brief music-listening task compare? What do you feel are some of the specific advantages? What do you feel are some of the specific disadvantages?
- Can you foresee any challenges when providing this to other healthcare staff?
  - Prompts: such as needing to encouraging them to access support for themselves as well as supporting other people? Making people aware that they are experiencing intrusive memories, and that something might be available to help them? Supporting people to find the time to use the intervention?
  - If so, how do you think these challenges could be addressed?
- How would you want to be offered this technology? Who would you want to manage the data or have access to it?

**Future usage**

- Would you use this intervention again in the future (outside of a research study)? Why or why not?
- Is there anything you feel is missing from the intervention or could be added to the intervention?
- Do you have any further comments to make on the study or the digital imagery competing task or brief music-listening task?

#### 12-to-24-week Interview Topic Guide

**Using the intervention over the course of the trial**

- Have you carried on using the intervention?
  - If YES, how often are you using it, and when do you use it / what prompts you to use it? Has how you use it changed over time at all?
  - If NO, why was that? Please give all the reasons that apply – there may be multiple e.g. they no longer had intrusive memories OR because they no longer wished to / felt engaged with the intervention i.e. too busy, tired of using it; OR just got out of habit / didn’t have time etc.
- How easy or difficult was it to incorporate this digital imagery competing task or brief music-listening task into your day?

*[Researcher note: Prior to interview, review participant’s study data. Are they still logging intrusive memories? At last available data, did intrusive memories for targeted memories seem to have reduced or not? Has the participant recorded and targeted multiple memories? Have they recorded and targeted any “new” memories? Does it seem that “old” memories have popped back? In general, does the intervention seemed to have worked for the participant, or not? ] [Tailor questions below to explore the pattern of study data you observe]*

- Have you experienced a change in the number of intrusive memories you are experiencing (We noticed your memories seems to have gone down / you stopped logging memories / etc)?
  - If yes, how has this affected how you day-to-day and at work?
  - If no (i.e., still having intrusive memories), can you tell me more about that? Did the intervention seem to not be effective? (Why, do you think?) Was there a reason that you stopped logging these intrusive memories / stopped using the intervention?

For intervention ppts. Only:

How many different memories did you target? Was this all of them? Did you reach a stage where there are no more remaining intrusive memories?

- If not, how do you feel about continuing or stopping using the intervention?

What shaped decisions around how many to target?

- Did they stop troubling you? What was the criteria for you to stop targeting them?
- How could we improve the intervention to make it easier to use or more effective over longer-term use, over weeks or months (or even years) of use?
- Have you started experiencing any new intrusive memories since you started taking part in the trial, perhaps as a result of difficult experiences at work?
  - If so, have you used the intervention to help try and reduce them?
  - If YES, how did that go? Can you share the process, and how you felt about using it for new memories?
  - If NO, why was that? There may be multiple reasons, so please give the range of reasons as it’s helpful for us to understand.

**Future usage**

- Would you use this intervention again in the future, outside of a research study?
  - [If yes or maybe] What would be your main motivation for using the intervention again in future? Do you think you would be primarily motivated by improvement in your own health and wellbeing, or by improvements in how you are doing at work or ability to deliver quality care, or something else?
  - [If no] Why not?
- What do you think would most motivate your colleagues to try the intervention, if they were experiencing intrusive memories? Would it primarily be improvement in their own health and wellbeing, or improvements in how they feel at work and in their ability to deliver care, or something else?
- Would you prefer to access this intervention through work (e.g., provided by your employer) or outside of work (e.g., something you decide to do yourself)? Do you think your colleagues would feel similarly?
- How comfortable would you feel if you were contacted about this intervention through your workplace and it was provided through work (e.g., through a Wellbeing Hub, or as a standard part of the staff training and appraisal process)?
  - Would this pose any difficulties for yourself or your colleagues? If so, are there ways in which we could try to overcome these difficulties?
- Do you have other thoughts on how the intervention could be made available through the workplace … for example, thinking about any other staff wellbeing services or initiatives that you’ve been aware of? What do you think has been successful, or not, and why?
- Do you think the intervention would be taken seriously by NHS staff experiencing intrusive memories, or is there a risk that it will be perceived as not very important to them, ‘just another wellbeing tool’, or just a game? What could help the intervention be recognised as important and useful?
- If you had to guess, how many NHS staff who have intrusive memories would be interested in trying this intervention? What percentage – e.g., 25%, 50%, 75%?

*(The following questions are optional and only to be asked if not addressed in earlier parts of the interview)*

- Are there any aspects of the digital imagery competing task or brief music-listening task that would put you off using it again in the future?
- Are there any aspects of the digital imagery competing task or brief music-listening task that make you more likely to use it again in the future?
- Would you recommend the study to any of your colleagues or friends? If so, what are the features that you would highlight? If not, what reasons would you give them?
- Do you think an approach like this to prevent the impact of trauma that arises in the job would be welcomed in the NHS, in relation to mental wellbeing? So, by managers and decision-makers, and in the workplace more generally? Why or why not? If not, what do you think would help to overcome these barriers?
